# Comprehensive Comparison of a Genotype-Forward vs. Phenotype-Forward Approach for Monogenic Cardiovascular Diseases in the UK Biobank Cohort

**DOI:** 10.1101/2025.09.11.25335614

**Authors:** Megan E. Ramaker, Kristin M. Corey, Jessica A. Regan, Sara Coles, Jawan W. Abdulrahim, Kalyani Kottilil, Navid Nafissi, Kaitlyn Amos, Meghan Mac Neal, Lydia Coulter Kwee, Senthil Selvaraj, Svati H. Shah

## Abstract

**Background:** Monogenic cardiovascular diseases (MCVD) remain substantially under-diagnosed, and thus interest is shifting from a phenotype-forward approach towards a genotype-first strategy that applies to population-wide genomic screening. We evaluated the prevalence of pathogenic and predicted pathogenic MCVD variants and assessed disease expression in the population-based UK Biobank (UKB) cohort to determine whether yield is greater in phenotype-versus genotype-forward approach.

**Methods:** Pathogenic and likely pathogenic (P/LP) variants, and variants of uncertain significance (VUS) predicted to be pathogenic (pp-VUS) in 47 MCVD genes were assessed. In the genotype-forward approach, UKB participants with P/LPs or pp-VUS were assessed for symptoms of disease using electronic health record (EHR) interrogation, emulating a clinical approach to population-based screening. In the phenotype-forward approach, participants with stricter EHR-based evidence of disease expression were identified (emulating current clinical care) and the presence of P/LPs and pp-VUS was determined.

**Results:** Following QC, 467,850 participants were included. Overall, 1 in 125 participants in UKB carry an MCVD P/LP. In the genotype-forward approach, 3,709 (0.79%) participants carried an MCVD P/LP and 42.1% of these had symptoms of the associated MCVD; another 29,269 (6.3%) carried a pp-VUS with 21.5% of these having symptoms of disease. In the phenotype-forward approach, 62,488 (13.4%) expressed an MCVD using strict evidence of disease expression and of these individuals 1% carried an associated P/LP variant and 2.4% carried an associated pp-VUS.

**Conclusion:** We show here that a genotype-forward approach leads to a 2.5 times higher diagnostic yield (3.6 times higher when considering pp-VUS) compared with the phenotype-forward approach, which missed 936 P/LP carriers with disease symptoms. While cost, access and ethics need to be considered, these results support expanded population-based genetic screening to improve diagnosis of potentially treatable MCVD.

## Introduction

Monogenic cardiovascular diseases (MCVD) encompass heterogeneous diseases that increase risk for severe cardiac complications including sudden death (1–3). The current clinical paradigm for evaluating MCVD relies on a “phenotype-forward” approach, where patients with symptoms of disease or family history are referred for clinical evaluation and genetic testing. Increased availability of preventive treatments and the underdiagnosis of MCVD highlight the need for better identification of individuals with MCVD-related disease expression and those at elevated risk due to presence of an MCVD genetic variant (4,5). However, most studies to-date characterizing genetic explanation of disease in participants with suspected MCVD is performed in cohorts with high suspicion for disease biasing the understanding of the yield of genetic screening (6).

The increased accessibility and decreasing costs of genetic sequencing have led to increased genetic sequencing in ostensibly healthy, population-based cohorts, which is beginning to reveal the prevalence of undiagnosed monogenic disease (5,7–10). Analysis of these populations could reveal the clinical utility of a genotype-forward approach to assessing for monogenic disease. For example, in the Cardiovascular Catheterization Genetics (CATHGEN) cohort where whole exome sequencing (WES) was performed in ∼8,500 patients being evaluated for cardiovascular disease, 4.5% carried an MCVD pathogenic (P) or likely pathogenic (LP) variant and had definitive or probable evidence of the associated disease, yet only 35% of those individuals had been given a clinical diagnosis (5). Such an approach has also been tested in population-based cohorts further revealing the rate of underdiagnosis of genetic diseases at a broader scale. Within the *BioMe* clinical care biobank, less than 2% of pathogenic and loss of function variant carriers had previously been diagnosed with the associated genetic disorder (11). By contrast, among the 3,350 eMERGE III participants who underwent genetic testing because their clinical presentation suggested an MCVD, only 1.9% were found to carry a P/LP variant (8). Early genetic testing has been shown to facilitate appropriate disease management including increased clinical monitoring and earlier preventative therapeutic strategies that can ultimately save lives (12). However, large studies have not directly compared a genotype- vs. phenotype-forward approach to MCVD in the same population.

We leveraged the UK Biobank (UKB) population-based cohort to determine whether the yield of a genotype-forward approach is greater than phenotype-forward approach for MCVD evaluation. We assessed the prevalence of P/LP variation and expressivity of disease at a population-scale, while additionally evaluating the yield when incorporating variants of uncertain significance predicted to be pathogenic (pp-VUS).

## Methods

### Study Population

The UK Biobank (UKB) study is a prospective study of 502,493 participants aged 40-69 recruited in 2006-2010 at 22 assessment centers from across the United Kingdom (National Research Ethics Service, 11/NW/0382) (13). Data about lifestyle, physical measures and self-reported medical conditions as well as biospecimens were collected on enrollment. Primary care records and hospital records were also obtained including International Classification of Diseases, 9^th^ and 10^th^ revisions (ICD-9 and ICD-10) diagnostic codes and OPCS-4 operation codes. A subset of participants underwent cardiac magnetic resonance imaging (CMR) (n=38,878), 12-lead ECG (n=77,576) and WES (n=469,686).

### Whole Exome Sequencing and Variant Calling

DNA extraction from whole blood and WES was performed by Regeneron as previously described (13). For this study, multi-sample project-level VCF (pVCF) files generated using GLnexus were utilized (14). Participants were excluded for sex mismatch, high rates of heterozygosity, sex chromosome aneuploidy, or if a participant had disenrolled from the study since the time of sequencing, leaving 467,850 participants retained for downstream analysis. Genetic variants were filtered by sequencing depth (≤10) and quality (≤20) using VCFtools (15). There was no exclusion for variant type, thus multiallelic sites, single nucleotide variants and indels were included. For genetic ancestry assessment, we utilized the UKB-provided ‘Genetic ethnic group’ field (22006), which indicates a ‘1’ if participants self-identified as ‘White British’ and have a similar genetic ancestry based on genetic principal components (Caucasian) and missing if participants do not meet these criteria.

### Monogenic Cardiovascular Disease Gene Prioritization and Variant Annotation

Forty-seven genes were selected based on inclusion in genetic testing panels and literature review for established association with the following MCVDs: cardiomyopathies (CM), hereditary transthyretin amyloidosis (hATTR), familial hypercholesterolemia (FH), arrhythmia disorders, and aortopathies/connective tissue disorders (Supplemental Table 1). ClinVar was used to identify pathogenic (P), likely pathogenic (LP), likely benign (LB), benign (B), conflicting variants and variants of uncertain significance (VUS) in these genes (16). Annotation for variants was downloaded from ClinVar on December 20^th^, 2023, and subset to the 47 genes using Bedtools V 2.30.0 (17). Ensemble variant effect predictor (VEP; version 106) and CADD version 1.6 were used for variant annotation (18,19).

### Confirmation of Pathogenic and Likely Pathogenic Variants

A semi-automated workflow previously established by our group was conducted using American College of Medical Genetics and Association for Molecular Pathology (ACMG/AMP) review based on the 2015 updated guidelines to determine if a previously ClinVar-defined P/LP variant was P or LP (20,21). Briefly, ACMG/AMP review was automated for criteria obtained from publicly available databases including: loss of function (PVS1), same amino acid change as a another P/LP variant (PS1), within a mutational hotspot or critical domain (PM1), absent large population databases (PM2), protein length changing (PM4), results in a different amino acid change in the same position of another P/LP (PM5), and *in silico* annotations for the variant (PP3). Criteria that could not be automated and were thus manually assessed included: functional studies (PS3) or case-control studies (PS4), cosegregation with disease (PP1), and reputable source reporting as P/LP (PP5). The following criteria could not be included in our ACMG/AMP review: whether the variant was *de novo* in a patient without a family history (PS2), in *trans* with a P/LP variant in recessive disorders (PM3), assumed to be *de novo* (PM6), missense variant in a gene with low rate of benign variation (PP2) or phenotype or family history highly specific for disease (PP4).

### Variant of Uncertain Significance (VUS) Pathogenicity Prediction

We previously developed a CVD-specific machine learning algorithm, cardiovascular disease pathogenicity predictor (CVD-PP), that prioritizes VUS, enabling discrimination of VUS that are more likely to be P/LP vs. B/LB variants (21). VUS were defined for this study as either being 1) present in UKB but absent in ClinVar or 2) classified as conflicting or uncertain in ClinVar. In brief, CVD-PP is a random forest algorithm that uses gnomAD allele frequency, Maximum Entropy Scan difference (22), CADD PHRED score (19), distance to the nearest ClinVar P/LP and GTEx cardiac (left ventricle and aorta) exon-level expression (23) as predictors of variant pathogenicity. The previously-defined CVD-PP cut-off for pathogenicity of 0.56 was utilized to classify VUS as “predicted pathogenic” (i.e. P/LP) vs “predicted benign” (i.e. B/LB). VUS that were predicted P/LP by the CVD-PP algorithm were retained for downstream analysis.

### Monogenic Cardiovascular Disease Phenotyping

Clinical disease phenotyping used all available ICD-9, ICD-10, OPSC-4, and self-report codes, along with EKG, CMR features and laboratory values from the EHR. All disease-specific codes used are within Supplemental Tables 2-6. Different criteria were used for the genotype- vs. phenotype-forward approach to simulate respective clinical workflows. For the genotype-forward approach, participants found to carry a P/LP or pp-VUS would be suspected of having the related MCVD (and thus have further clinical evaluation) even with relatively non-specific criteria or symptoms (i.e. carriage of a familial cardiomyopathy P/LP and presence of shortness of breath). For the phenotype-forward approach, as is done in current practice, participants with evidence of MCVD based on stricter criteria (i.e. cardiomyopathy without coronary artery disease) are referred for genetic testing and considered to have the related MCVD if found to carry a respective P/LP variant.

Phenotypic classifications were defined and grouped within each category by expert manual review. Similar phenotype criteria for CM (dilated cardiomyopathy [DCM] and hypertrophic cardiomyopathy [HCM]; Supplemental Table 2). For aortopathies/connective tissue disorders, in addition to common phenotypes across all disease subtypes (i.e. aortic aneurysm/dissection), criteria unique to subtypes were also included (i.e. the ICD code for Marfan’s syndrome [Q74]; Supplemental Table 5). Similarly for arrhythmia disorders, broader arrhythmia codes were utilized for all disorders in addition to more specific criteria for disease subtypes: Brugada syndrome (BrS), long QT syndrome (LQTS), atrial fibrillation (AFib) and arrhythmogenic right ventricular cardiomyopathy (ARVC; Supplemental Table 6).

## Results

### Overall Prevalence of MCVD Variants in UKB

In the genotype-forward approach, we first evaluated the prevalence of P/LP variants in MCVDs within the UKB (Figure 1). In 47 MCVD genes, 146,358 variants were identified, of which 821 (0.6%) were annotated by ClinVar as P/LP variants (Figure 2a) and 719 (87.6%) variants present in 43 of the 47 MCVD genes were confirmed via ACMG/AMP review. We additionally assessed for presence of VUS predicted to be pathogenic (pp-VUS). Most of the 146,358 variants (94.7%) were VUS (absent in ClinVar: n=132,155 or ClinVar conflicting or uncertain: n=6,467). *In silico* prediction of variant pathogenicity was performed using CVD-PP to further stratify these VUS, resulting in 9,207 pp-VUS (6.6%) being predicted pathogenic (Figure 2b).

**Figure 1.**
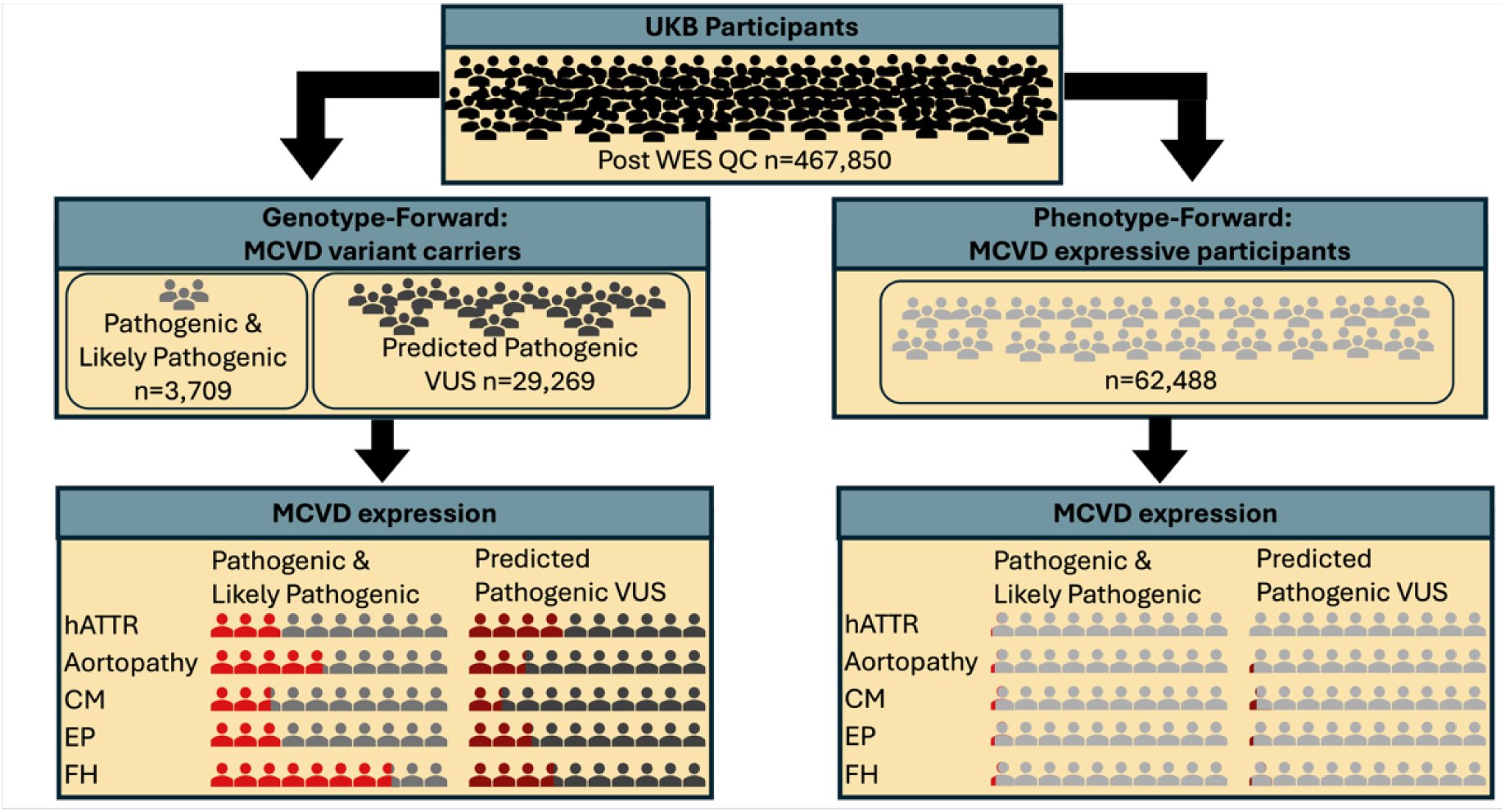
Design of the Study. UK Biobank participants with WES data that passed QC (n=467,850) were filtered for pathogenic and likely pathogenic (P/LP, as defined in the ClinVar database and ACMG-AMP reviewed by clinicians) as well as predicted pathogenic variants (pp-VUS, as defined by a previously published model (21)) in 47 monogenic cardiovascular disease (MCVD) genes; carriers were then assessed using more liberal phenotype criteria for evaluation of expression of the associated MCVD (genotype-forward approach, left). In a phenotype-forward approach (right), participants were assessed for evidence of MCVD using more strict phenotypic criteria followed by filtering for P/LP and pp-VUS.

**Figure 2.**
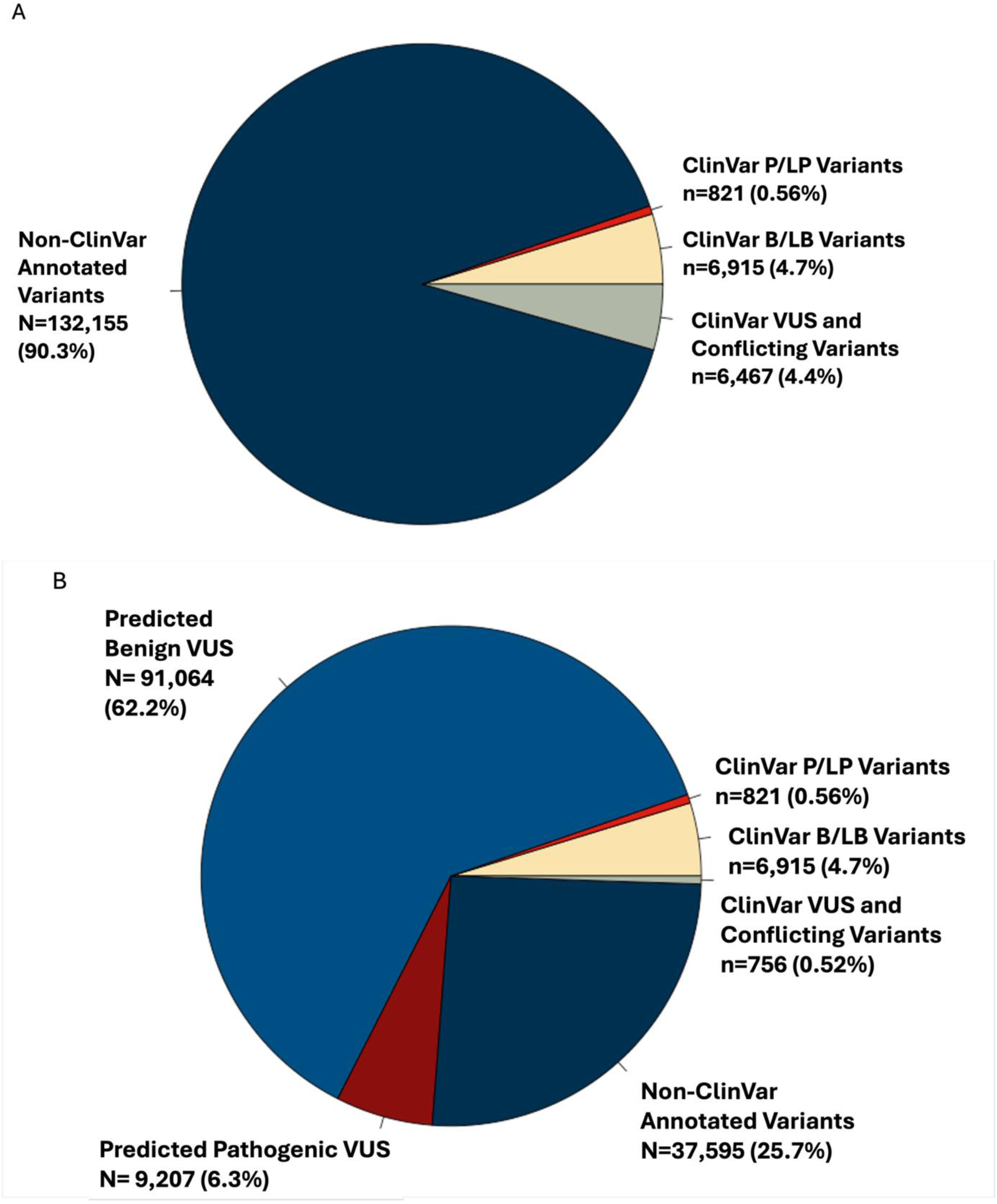
Categories of MCVD Genetic Variants Identified. A pie chart demonstrating the distribution of 146,358 identified variants in 47 MCVD genes, with proportion by clinical classification in the ClinVar database, the majority being not present in the ClinVar database (panel A); and distribution after application of the Cardiovascular Pathogenicity Predictor model on the 138,622 VUS, resulting in classification of the majority of variants of uncertain significance into predicted P/LP or predicted B/LB (panel B). P/LP = Pathogenic and Likely Pathogenic. B/LB = Benign and Likely Benign. VUS = Variant of Uncertain Significance.

### Prevalence of P/LP Genetic Variants by MCVD

We identified 436 unique P/LPs within 36 of 47 MCVD genes present in 467,850 UKB participants with WES following QC (Supplemental Table 7). Overall, 3,709 participants (0.79%) harbored at least one ACMG/AMP confirmed P/LP variant (693 variants across 36 MCVD genes; Table 1). The prevalence of P/LPs varied by MCVD, where 1,202 (0.26% of total population) participants carried a CM P/LP; 437 (0.09%) carried an hATTR P/LP; 169 (0.04%) carried an aortopathy/connective tissue disease P/LP; 895 (0.19%) carried an arrhythmia P/LP; and 1,003 (0.21%) carried an FH P/LP (Figure 3, Table 2).

**Figure 3.**
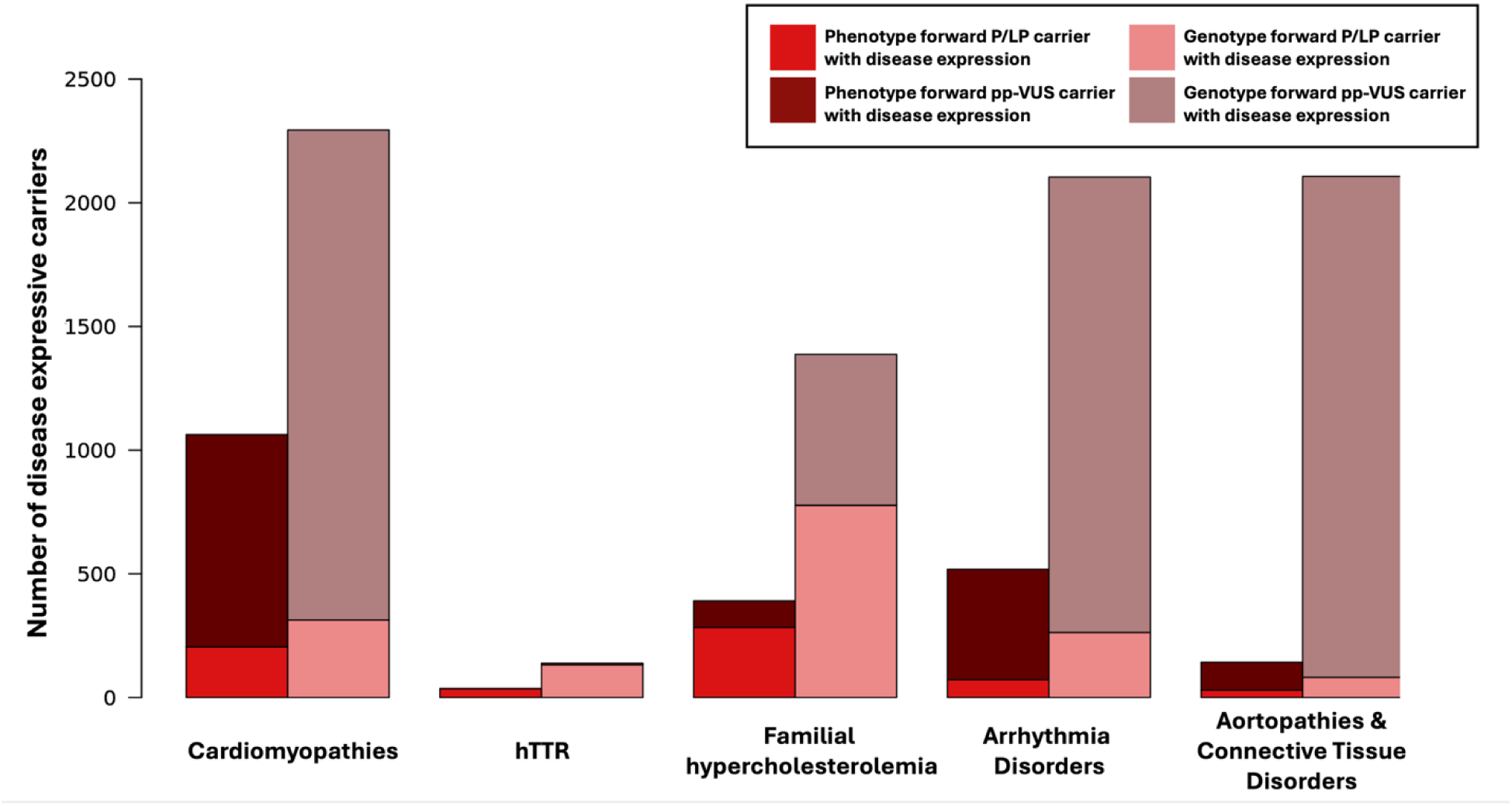
Number of pathogenic and likely pathogenic as well as predicted pathogenic variant carriers with evidence of disease expression by monogenic cardiovascular disease category. A bar graph is displayed demonstrating the number of pathogenic and likely pathogenic (P/LP) or predicted pathogenic variant of uncertain significance (pp-VUS) carriers with evidence of disease expression (Y-axis), for the phenotype-forward (light [P/LP] and dark [pp-VUS] red bars) and genotype-forward (light [P/LP] and dark [pp-VUS] pink bars) approaches, demonstrating that a genotype-forward approach identified more P/LP carriers with disease expression across MCVD disease categories.

**Table 1.** Basic demographics of carriers of pathogenic/likely pathogenic (P/LP) and predicted pathogenic (pp-VUS) variants.

|  |  |  |  |  |
| --- | --- | --- | --- | --- |
| **Number of participants** | 3,709 (0.79%) | 1,562 (42.1%) | 29,269 (6.3%) | 6,287 (21.5%) |
| **Age (mean +/- SD)** | 56.0 (+/-8.2) | 57.8 (+/-7.9) | 56.3 (+/-8.1) | 59.5 (+/-7.3) |
| **Percent female sex (N)** | 56.3% (2,090) | 52% (812) | 53.7% (15,729) | 45.4% (2,854) |
| **Percent Caucasian Genetic Ethnic Grouping (N)** | 76.0% (2,819) | 78.9% (1,232) | 81.8% (23,954) | 83.4% (5,243) |

**Table 2.** Numbers and proportion of P/LP and pp-VUS carriers with evidence of disease expression by MCVD category, genotype-forward approach.

| **Disease category** | **Pathogenic and Likely Pathogenic Variant Carriers (% from WES)** | **Disease Expressive Pathogenic and Likely Pathogenic Variant Carriers (% of carriers)** | **Predicted Pathogenic Variant Carriers (% of WES)** | **Disease Expressive Predicted Pathogenic Variant Carriers (% of WES)** |
| --- | --- | --- | --- | --- |
| **Familial Hypercholesterolemia** | 1,003 (0.21%) | 775 (77.3%) | 1,627 (0.35%) | 606 (37.2%) |
| **Cardiomyopathies** | 1202 (0.26%) | 313 (26.04%) | 13,245 (2.8%) | 1,943 (14.7%) |
| **hATTR** | 437 (0.09%) | 132 (30.2%) | 18 (0.004%) | 7 (38.9%) |
| **Arrhythmia** | 895 (0.19%) | 264 (29.5%) | 6740 (1.4%) | 1,859 (27.6%) |
| **Aortopathies/connective tissue disorders** | 169 (0.04%) | 82 (48.5%) | 8,389 (1.8%) | 2,001 (23.9%) |

Most P/LP carriers had only one MCVD P/LP variant, with only 13 (0.003% of the total cohort) participants carrying two P/LP variants (Supplemental Table 8). Two of these participants were compound heterozygous for a P/LP within the same gene and the remainder carried P/LPs in different genes. The most commonly occurring combination in these carriers was a P/LP in a CM gene along with an ARVC-associated P/LP (Supplemental Figure 1).

### Prevalence of pp-VUS

There were 3,172 unique pp-VUS present in 45 of 47 MCVD genes following WES QC, annotation and pathogenicity prediction with CVD-PP (Supplemental Table 9). Overall, 29,269 participants (6.3%) without a known P/LP carried at least one of 7,736 pp-VUS across 45 genes (Figure 1, Table 1). The number of pp-VUS carriers varied across MCVDs, where 13,245 participants (2.8% of total population) carried a CM pp-VUS; 18 (0.004%) participants carried an hATTR pp-VUS; 8,389 (1.8%) participants carried an aortopathy/connective tissue pp-VUS; 6,740 participants (1.4%) carried an arrhythmia pp-VUS; and 1,627 participants (0.35%) carried an FH pp-VUS (Figure 3, Table 2). Of pp-VUS carriers, 999 (3.4%) had two pp-VUS identified (Supplemental Table 10). Of these, the most common combination was two pp-VUS within a CM gene (Supplemental Figure 2).

### Genotype-forward Approach to MCVD: Overall Disease Expression in Carriers

In a genotype-forward approach to identifying MCVD, 1,562 (42.1%) P/LP carriers demonstrated symptoms of the associated MCVD using more liberal criteria. Of 29,269 MCVD pp-VUS carriers, 6,287 (21.5%) had symptoms of the associated MCVD (Table 1). The gene with the highest proportion of P/LP carriers with disease symptoms was *LDLR,* with 83.5% showing evidence of disease (Figure 4a, Supplemental Table 11). There were five MCVD genes for which no participant harboring a P/LP variant demonstrated disease symptoms: *TPM1, MYH11, KCNE2, KCNE1,* and *DSC2*. Similarly to P/LP results, the gene with the highest proportion of pp-VUS carriers with evidence of gene expression was *LDLR,* with 52.3% of carriers showing evidence of disease expression (Figure 4b, Supplemental Table 12). Thus, using a genotype-forward approach, 1,562 participants (0.33% of total population) both had expression of disease and carried a P/LP variant (increasing to 6,287 [1.3% of total population] when including pp-VUS).

**Figure 4.**
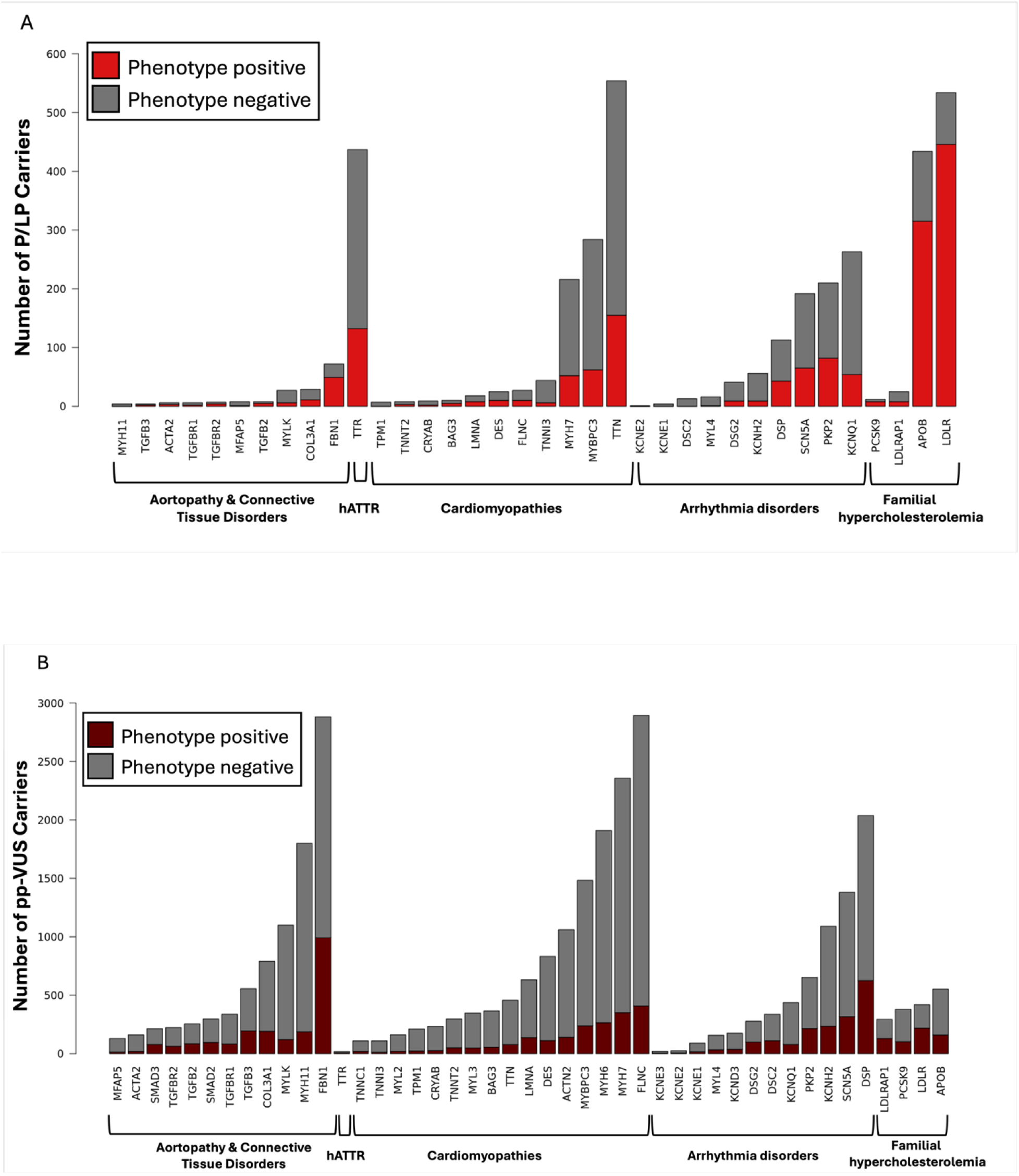
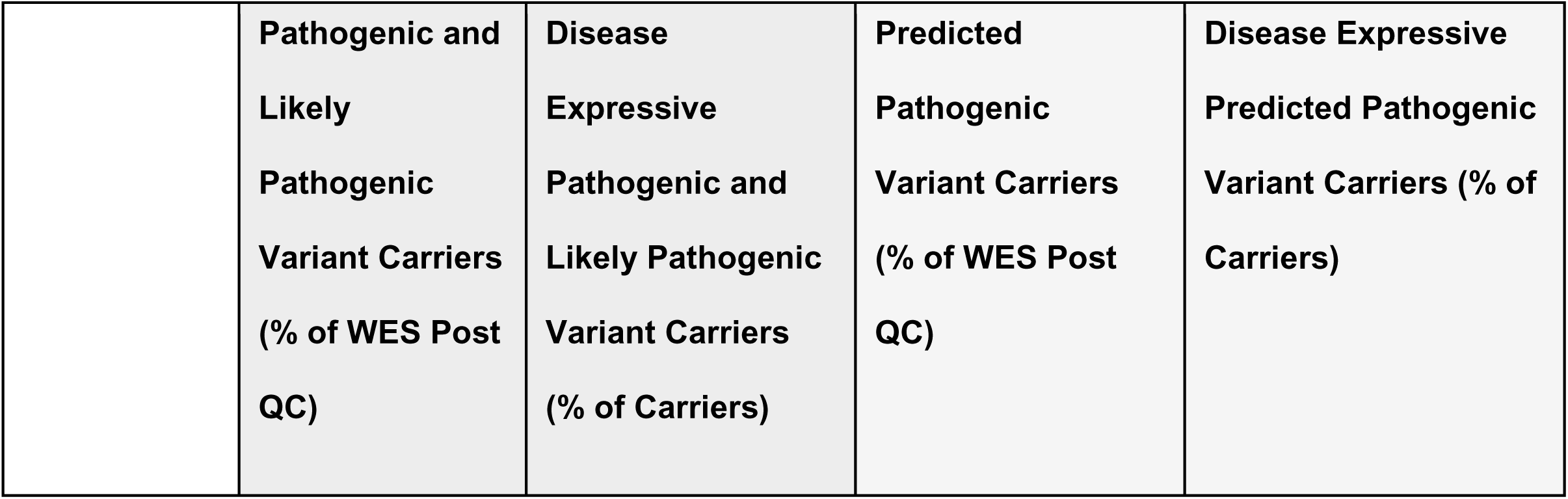
Number of participants carrying a pathogenic and likely pathogenic variant, as well as a predicted pathogenic variant by gene. Displayed are bar graphs demonstrating the number of participants carrying a pathogenic or likely pathogenic (P/L) (panel A) or predicted pathogenic variant of uncertain significance (pp-VUS; panel B).

### Genotype-forward Approach to MCVD: Disease-Specific Expression in Carriers

Of MCVDs, FH had the highest number of P/LP carriers with symptoms of disease (n=775; 0.17% of total population; Figure 3, Table 2). In FH genes, the percentage of carriers with disease symptoms ranged from 32% for *LDLRAP1* P/LP carriers to 83.5% for *LDLR* P/LP carriers (Figure 4a, Supplemental Table 11). Additionally, 1,627 participants (0.35% of the total population) had an FH pp-VUS, of whom 37.2% had evidence of disease symptoms (Figure 3, Table 2).

For CM, of 1,003 participants (0.26% of the total population) harboring a P/LP variant 26% expressed symptoms of disease (Figure 3, Table 2). In CM genes, the percentage of carriers with disease symptoms ranged from 0% for *TPM1* to 50% for *BAG3* (Figure 4a, Supplemental Table 11). Of the 13,245 participants (2.8% of the total population) who carried a pp-VUS in a CM-associated gene, 14.7% expressed CM symptoms (Figure 3, Table 2).

For hATTR specifically, there were 12 distinct *TTR* P/LP variants present in 437 participants, of whom 132 (30.2%) had evidence of disease symptoms (Supplemental Table 7). The P/LP variant most frequently observed was Val142Ile (historically Val121Ile) with 29.5% of Val142Ile carriers having evidence of disease symptoms. Three Val142Ile carriers were homozygous, all of whom had evidence of disease symptoms. The majority of Val142Ile carriers (97.3%) were not designated ‘Caucasian’ genetic ancestry by UKB (field ID 22006) and most (80.9%) self-reported Black or Caribbean ethnic background. Additionally, only 18 participants (0.004%) carried a *TTR* pp-VUS, mostly of UKB-determined Caucasian background (77.8%), of whom 38.9% had evidence of disease symptoms (Figure 4b, Supplemental Table 9).

For arrhythmia disorders, 895 participants (0.19% of total population) carried an arrhythmia-associated P/LP and 264 (29.5%) had evidence of disease symptoms (Figure 3, Table 2). In arrythmia genes, the percentage of carriers with disease symptoms ranged from 0% for *DSC2, KCNE1* and *KCNE2* P/LP carriers to 39% in *PKP2* P/LP carriers (Figure 4a, Supplemental Table 11). Amongst 6,740 participants (1.4% of total population) who carried at least one arrhythmia pp-VUS, 27.6% had evidence of disease symptoms (Figure 3, Table 2).

For aortopathy/connective tissue diseases, 169 participants (0.04% of total participants) carried one of 85 P/LPs and 48.5% had evidence of disease symptoms (Figure 3, Table 2). For aortopathy/connective tissue diseases genes, the percentage of carriers with evidence of disease symptoms ranged from 0% for *MYH11* P/LP carriers to 68.1% for *FBN1* P/LP carriers (Figure 4a, Supplemental Table 11). Amongst the 8,389 participants (1.8% of total population) who carried an aortopathy/connective tissue disease pp-VUS, 23.9% had evidence of disease symptoms (Figure 3, Table 2).

### Genotype-Forward Approach: Disease Expression in Participants with Two P/LP

Of the 13 participants (0.003 % of total population) who carried two P/LPs in any MCVD gene 61.5% expressed symptoms of associated disease for either P/LP they carried, 2 of whom were compound heterozygous for the same gene (Supplemental Table 8, Supplemental Figure 1). In participants with more than one pp-VUS, 292 (29.2%) expressed associated disease symptoms with at least one of the pp-VUS they harbored (Supplemental Table 10, Supplemental Figure 2). In these participants, the most frequently observed combination was two pp-VUS within CM-associated genes (n=215) of whom 17.7% had evidence of disease expression.

### Phenotype-Forward Approach: Yield of Genetic Diagnosis in Participants Suspected to Have MCVD

In the phenotype-forward approach, 62,488 participants (13.3% of total population) had phenotypic evidence of at least one MCVD disease using more strict criteria (Table 3). The most prevalent MCVD in the UKB using these criteria was FH (7.3%), followed by arrhythmia disorders (5.8%), hATTR (4.7%), CM (5.4%), and aortopathies/connective tissue disorders (1.1%) (Figure 3, Table 3). Of these participants, 1% carried a P/LP in the associated MCVD gene and an additional 2.4% carried a pp-VUS in the associated MCVD gene (Figure 3, Table 3). Thus, using a phenotype-forward approach, 626 participants (0.13% of total population) both had expression of disease using strict criteria and carried a P/LP variant (increasing to 2,152 [0.46% of total population] when including pp-VUS). Observed codes in the phenotype-forward vs the genotype-forward in carriers are listed in Supplemental Tables 15-19.

**Table 3.** Phenotype-forward monogenic cardiovascular disease expression and genetic variant carrier status for both pathogenic and predicted pathogenic variants by disease grouping.

| **Disease category** | **Number of phenotype positive participants using strict criteria (% from WES)** | **Number of phenotype and P/LP positive participants (% from phenotype positive)** | **Number of phenotype and predicted pathogenic VUS positive participants (% from phenotype positive)** |
| --- | --- | --- | --- |
| **Familial Hypercholesterolemia** | 34,145 (7.3%) | 283 (0.83%) | 108 (0.32%) |
| **Cardiomyopathies** | 25,404 (5.4%) | 205 (0.81%) | 845 (3.3%) |
| **hATTR** | 22,016 (4.7%) | 36 (0.16%) | 0 (0%) |
| **Arrhythmia** | 273,32 (5.8%) | 72 (0.26%) | 433 (1.6%) |
| **Aortopathies/connective**<br>**tissue disorders** | 4,953 (1.1%) | 30 (0.61%) | 111 (2.2%) |

#### Phenotype-Forward Approach: Yield of Genetic Diagnosis by MCVD Category

In analyses of each individual MCVD class in the phenotype-forward approach, 283 participants (0.06% of total population) were identified as having strict FH criteria and carried an FH-associated P/LP. Additionally, 108 (0.02%) had strict FH criteria and carried an FH-associated pp-VUS (Figure 3, Table 3). FH carriers identified through this approach had high LDL (>4.9 mmol/L; Supplemental Table 15, Supplemental Figure 3).

For CM, 205 participants (0.04% of total population) were identified as having strict CM criteria and carried a CM-associated P/LP, while 845 (0.18% of total population) had strict CM criteria and carried a CM-associated pp-VUS (Figure 3, Table 3, Supplemental Tables 13-14). CM variant carriers identified in this approach tended to have heart failure ICD codes, including Left ventricular failure (I501) and congestive heart failure (I500; Supplemental Table 16, Supplemental Figure 4). Additionally, for hATTR, 36 participants identified as having strict criteria for hATTR diagnosis (0.008% of total population) carried a *TTR* P/LP. These carriers tended to have ICD codes for heart failure (I50; Supplemental Table 17 and Supplemental Figure 5).

For arrhythmia disorders, 72 participants (0.02% of total population) had strict arrythmia disorder criteria and carried an arrythmia-associated P/LP variant, and 433 participants (0.09%) had strict arrythmia disorder criteria and carried an arrythmia-specific pp-VUS (Figure 3, Table 3, Supplemental Tables 13-14). Arrhythmia variant carriers identified in this approach tended to have heart failure ICD codes (I50) in additional to arrhythmia codes (ventricular tachycardia [I472] and cardiac arrest with successful resuscitation [I460]; Supplemental Table 18, Supplemental Figure 6).

For aortopathy/connective tissue disorders, 30 participants (0.006%) had strict aortopathy/connective tissue disorder criteria and carried an aortopathy/connective tissue disorder-associated P/LP and 111 participants (0.02%) carried an aortopathy/connective tissue disorder-associated pp-VUS (Figure 3, Table 3). Aortopathy/connective tissue variant carriers identified in this approach tended to have aneurysm ICD codes (I71; Supplemental Table 19, Supplemental Figure 7).

## Discussion

By leveraging a large, population-based cohort, ACMG/AMP review of ClinVar P/LP variants and inclusion of variants predicted to be pathogenic, we found that a genotype-forward approach had greater yield for identifying participants with an MCVD compared with the current clinical paradigm of a phenotype-forward approach. This work reveals the prevalence of MCVD P/LP variants in an ostensibly healthy, population-based cohort and improves clinical characterization of the MCVD-associated genetic landscape by interrogating disease symptoms and evidence of disease expression of MCVD-associated genetic variation, revealing both consistencies and differences from previous findings (8,24–27).

By using a genotype-forward approach, we identified that a relatively high proportion of participants (1 in 125) who carried a P/LP variant in an MCVD gene, highlighting the potential burden of MCVD risk in a non-disease selected general population. Among these participants, 42.1% express signs or symptoms of the related disease, equating to 0.33% of the population both harboring a P/LP variant and expressing phenotypic evidence of the related MCVD. In contrast, when we applied the phenotype-first strategy used currently in clinical practice, 13.4 % of participants met the stricter MCVD phenotypic criteria that would ordinarily trigger genetic sequencing. Only 1% of this subgroup carried a corresponding P/PL. Thus, 1,562 participants (0.33% of total population) who both harbored an MCVD P/LP variant and had evidence of MCVD disease expression were identified using the genotype-forward approach as compared with 626 participants (0.13% of total population) using the phenotype-forward approach (Figure 3, Table 3, Supplemental Tables 13-14).

As such, 936 participants (0.2% of total population) who had an MCVD P/LP variant and more liberal evidence of disease expression would have been missed by the phenotype-forward approach. These yields were increased to 6,287 participants (1.3% of total population) for genotype-forward approach, and 2,152 participants (0.46% of total population) for the phenotype-forward approach, when including VUS identified as predicted pathogenic (pp-VUS) using a validated cardiovascular-specific pathogenicity model (21). These differences are related to the different phenotyping criteria used in the two approaches, with the genotype-forward approach emulating a potential clinical approach where the presence of P/LP variant would prompt suspicion of MCVD even with less severe disease progression or with subclinical disease.

Our results also highlight the complexity and heterogeneity of disease expression of MCVDs. In participants harboring an associated P/LP using the genotype-forward approach criteria, evidence of disease symptoms ranged from 27% in CM, 30% in arrhythmia disorders, 49% in aortopathies/connective tissue disorders and as high as 77.3% in FH in participants harboring an associated P/LP. We also show that 37% of P/LP variants associated with disease expression were observed in one participant (unique), further revealing the complex genetic landscape of MCVD. These results highlight the burden of MCVDs in the general population, the variability in disease expression and presentation among carriers of well-described P/LPs in MCVD-associated genes, and lastly the importance of VUS interrogation. As both precision health research and clinical efforts move towards more population-based genetic sequencing, these results provide both important considerations that should be made as well as a reference for variant expressivity across MCVD genes.

When compared to other studies, we find both similarities and differences in MCVD P/LP prevalence and evidence of disease expression with prior studies (24,28–31). In previous analyses of FH in the UKB, differences in observed prevalence of FH P/LP variants are likely due to the differing strategies in which pathogenicity is defined for variants. Expert review reporting previously reported a slightly higher prevalence at 0.27% (28) which was even higher in another study using an overall minor allele frequency cut off which revealed a 0.35% prevalence (29) compared to what we show (0.21%) using semi-automated ACMG review of ClinVar defined P/LPs.

When comparing prevalence of CM-associated P/LPs, Bourfiss et al identified a higher prevalence of P/LPs (0.73%) in overlapping CM-associated genes (*TPM1, TNNT2, BAG3, LMNA, DES, FLNC, TNNI3, MYH7, MYBPC3, TTN)* in a smaller release of the UKB WES data compared to what we find in this current analysis for the same gene set (0.25%) (30). While individual gene-level prevalence was not performed, Bourfiss et al report a much higher diagnosis rate of CM and/or heart failure in carriers of CM-associated P/LPs compared to matched controls and we similarly find a high prevalence of CM and heart failure diagnosis codes amongst CM-associated P/LP carriers in our analysis.

In comparison to a different analysis of LQTS-associated P/LP prevalence in the Trans-Omics for Precision Medicine (TOPMed) cohort, 160 (0.59%) P/LP carriers were identified across 10 LQTS-associated genes for which 5 overlap with the LQTS-associated genes assessed in this work (*KCNE1, KCNE2, KCNH2, KCNQ1,* and *SCN5A)*, where we identified a much lower prevalence of P/LP carriers (n=670; 0.14%) likely owing to the looser criteria for variant pathogenicity (ClinVar P/LP without ACMG/AMP confirmation or minor allele frequency < 0.01) (31). Lastly, for aortopathy/connective tissue disorders, we find concordant prevalence (0.02%) of *FBN1* P/LPs and disease expressivity (68.1%) with the estimated prevalence of disease between 0.02% and 0.01% (32–35).

This work highlights the potential utility and yield of a genotype-forward approach to population-based genetic sequencing and reveals individual MCVD P/LP prevalence and disease expression in variant carriers in a large, population-based study. Each MCVD phenotype criteria was determined by a respective clinician expert based on available codes in the UKB database. This approach helps overcome the limitation of using a large, population-based cohorts with EHR-based data, mostly consisting of billing codes that have known imprecision in capturing disease prevalence in participants (21,36–38). Further, our work highlights the burden of VUS while providing variant-specific results for VUS and variants absent from ClinVar predicted to be pathogenic, demonstrating the potential for increased yield at a population-level and expanding characterization of MCVD disease genes. The use of disease-agnostic *in silico* tools for variant prediction is only one criterion of many incorporated into ACMG/AMP criteria, but we note that our pp-VUS were identified using an MCVD-specific tool that has previously demonstrated high accuracy in identifying clinically significant variation (21). Lastly, our detailed variant specific results, provided in the supplementary information, may assist in variant classification and thus provide clinical utility.

We also note important limitations to our work. The UKB is a majority northern European ancestry cohort and thus does not allow for equal characterization of pathogenic variation in non-European participants, populations for which an expanded understanding is greatly needed. Furthermore, we reiterate the imprecision of using EHR data when assessing phenotypic expression of disease. It is widely accepted that EHR data are subject to several sources of bias, as they not only reflect patient health but also patients’ interactions with the healthcare system as well as healthcare processes (21,36–38). Additionally, the semi-automated ACMG/AMP review performed allowed for analysis of many P/LPs across 47 MCVD genes, however due to the nature of this project as a population-based study, we excluded some criteria that are helpful in determining pathogenicity of genetic variants (PS2, PM3, PM6, PP2 and PP4). Importantly, we also could not include a family history of disease in our workflow for phenotype-forward analysis, which is clinically used for referral for genetic testing and additional risk assessment. Finally, and perhaps most significantly, our study does not consider other important aspects necessary before a genotype-forward approach for population testing can be used more broadly. We did not evaluate cost considerations, access to genetic testing, ethical issues including the need for pre-test counseling and potential implications for downstream eligibility for life and other types of insurance not protected by the United States Genetic Non-Discrimination Act (GINA), the need for increased patient and health care provider genetics education, or appraisal of participant/patient preferences and attitudes around desire to receive this type of personal health information.

## Conclusions

Overall, our results suggest utility of an ostensibly healthy, population-based genetic sequencing approach and the need for continued and expanded evaluation of this methodology for diagnosing MCVD. Here we identified that in the UKB cohort, approximately 1 in 125 participants harbor P/LP variants in 47 MCVD genes, highlighting the relatively high prevalence in aggregate of these rare diseases. We show that a genotype-forwadr approach increases yield for MCVD compared to the widely-used phenotype-forward approach. Further, we find a high prevalence of VUS in MCVD and use a cardiac-specific machine learning tool (CVD-PP) to refine characterization of VUS.

## Abbreviations

MCVD: monogenic cardiovascular disease
P/LP: pathogenic/likely pathogenic
B/LB: benign/likely benign
VUS: variant of uncertain significance
pp-VUS: predicted pathogenic
VUS: CM, cardiomyopathy
DCM: dilated cardiomyopathy
HCM: hypertrophic cardiomyopathy
hATTR: hereditary amyloid transthyretin
FH: familial hypercholesterolemia
BrS: Brugada syndrome
LQTS: long QT syndrome
AFib: atrial fibrillation
ARVC: arrhythmogenic cardiomyopathy

## Web Resources

GTEx, https://www.gtexportal.org/home/index.html

ClinVar, https://www.clinicalgenome.org/data-sharing/clinvar/ UK Biobank Dataset, https://biobank.ctsu.ox.ac.uk

## Data Availability

UK Biobank data is available through the UK Biobank.

## Data Availability

UK Biobank data is available through the UK Biobank.

## Acknowledgements

The authors thank the UK Biobank participants for their participation in the study. This research has been conducted using the UK Biobank Resource under project number 48785.

## Author Contributions

M.E.R is responsible for conceptualization, data curation, formal analysis, software , investigation, methodology, and writing – original draft. K.M.C is responsible for data curation, formal analysis, methodology and investigation. J.A is responsible for conceptualization, data curation, and investigation. M.M.N is responsible for investigation. S.C, N.N, K.A, J.A.R and L.C.K are responsible for data curation. S.H.S is responsible for conceptualization, resources, supervision, and funding acquisition. All authors are responsible for writing – review and editing.

## Supplemental Figures

**Supplemental Figure 1.**
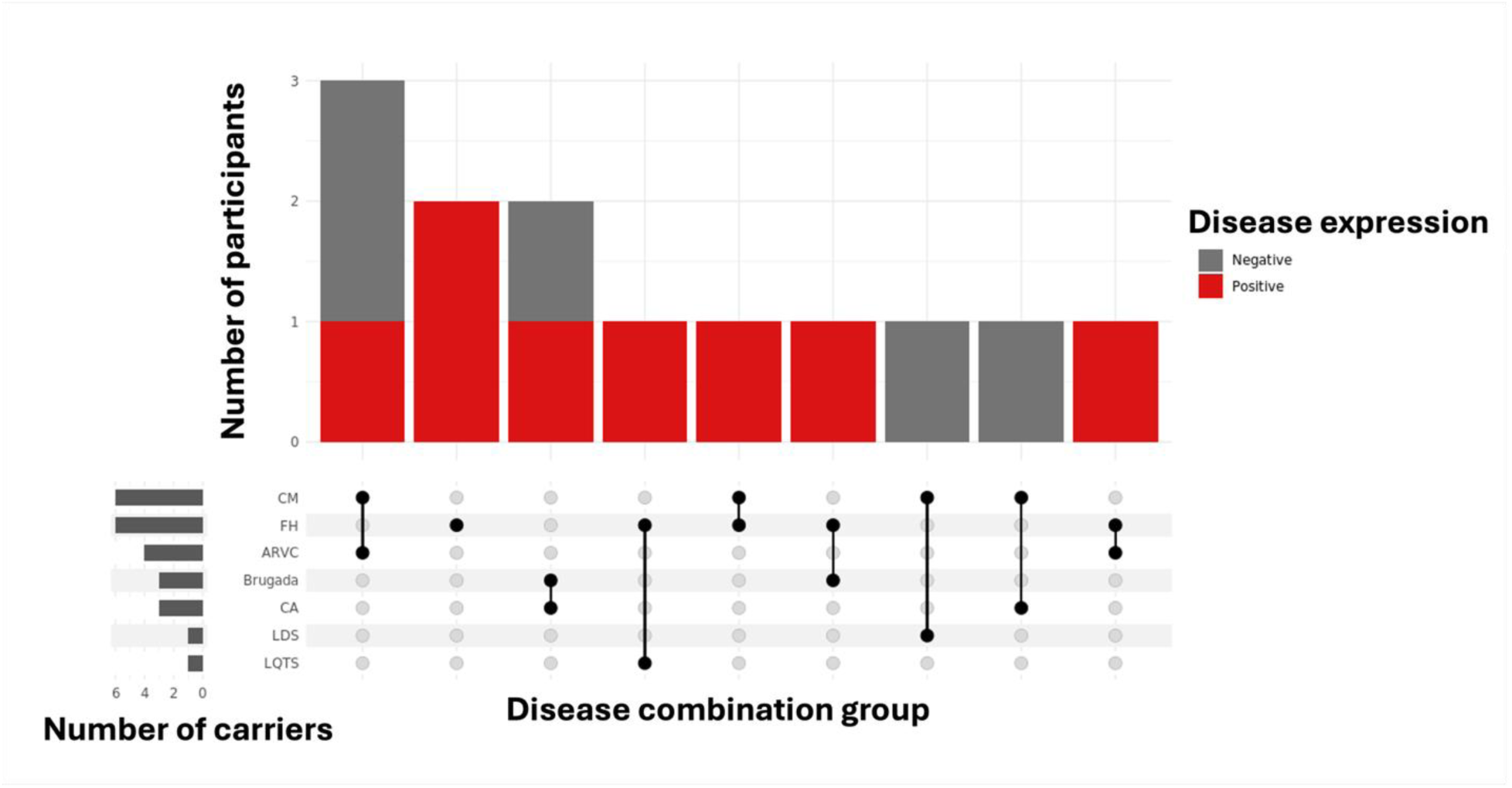
Upset plot of disease expressivity in carriers of two pathogenic or likely pathogenic variants by monogenic cardiovascular disease group.

**Supplemental Figure 2.**
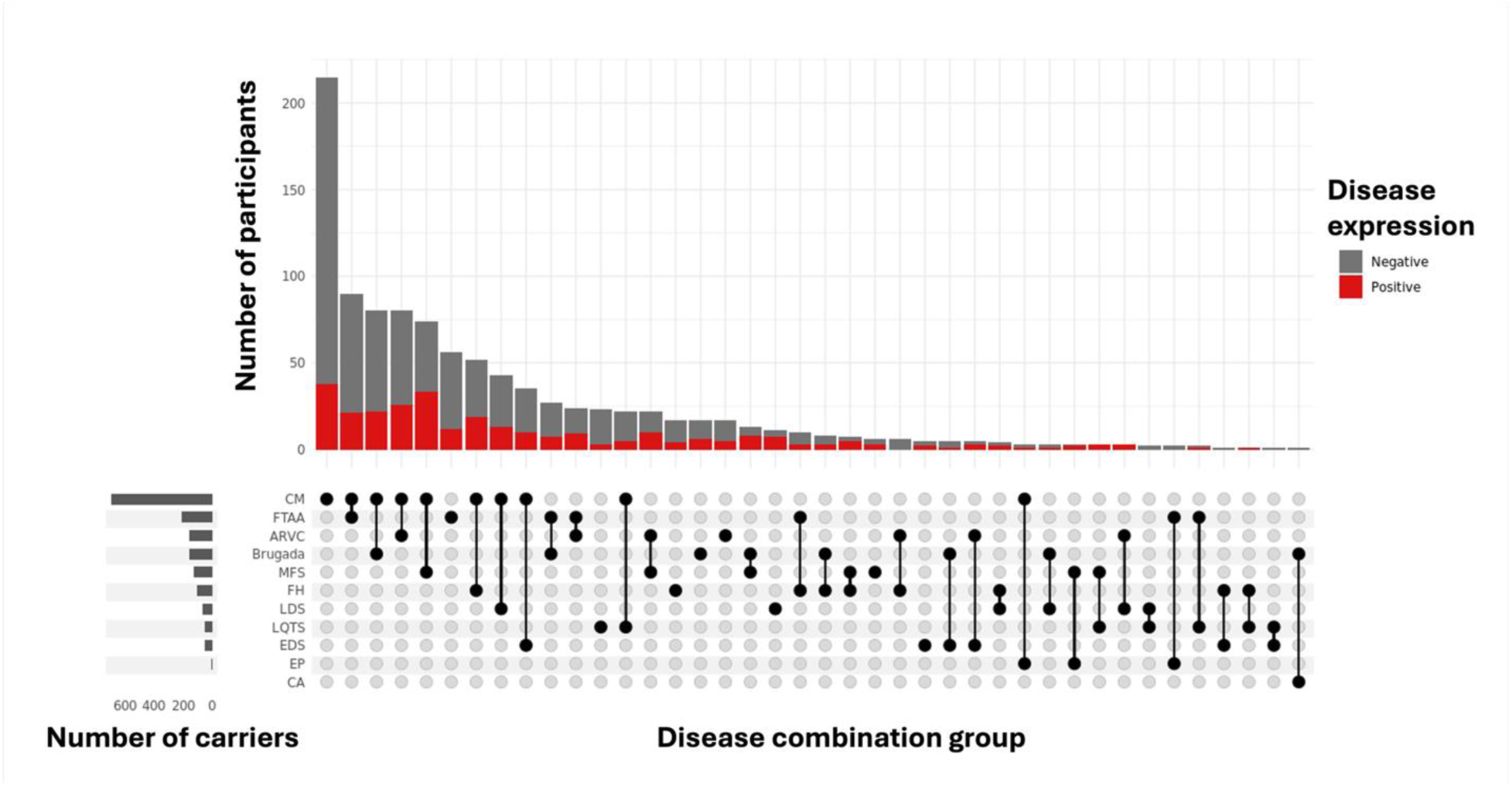
Upset plot of disease expressivity in carriers of two predicted pathogenic variants of uncertain significance by monogenic cardiovascular disease group.

**Supplemental Figure 3.**
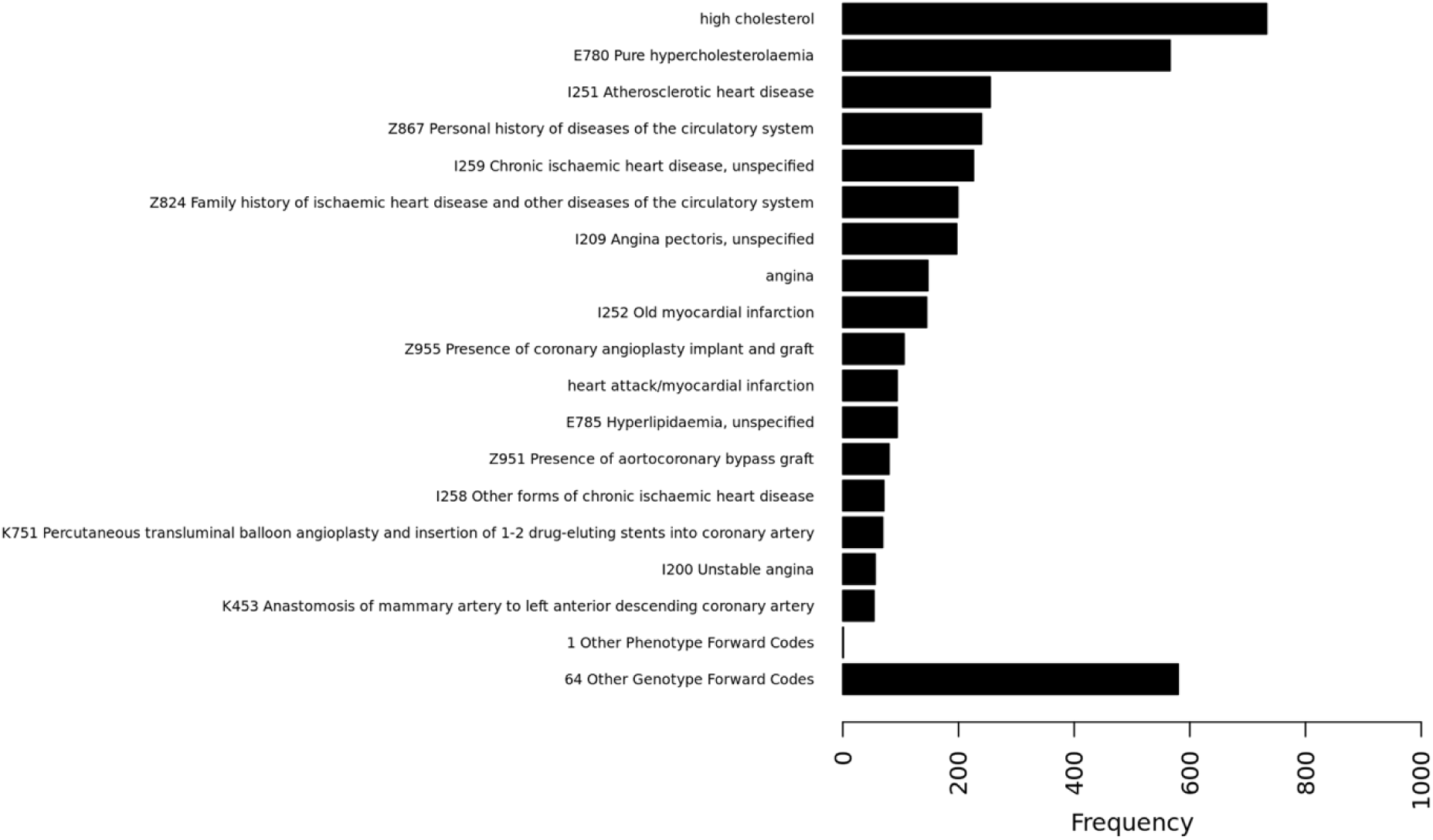
Familial hypercholesterolemia code frequencies observed in pathogenic or likely pathogenic or predicted pathogenic variant of uncertain significance carriers.

**Supplemental Figure 4.**
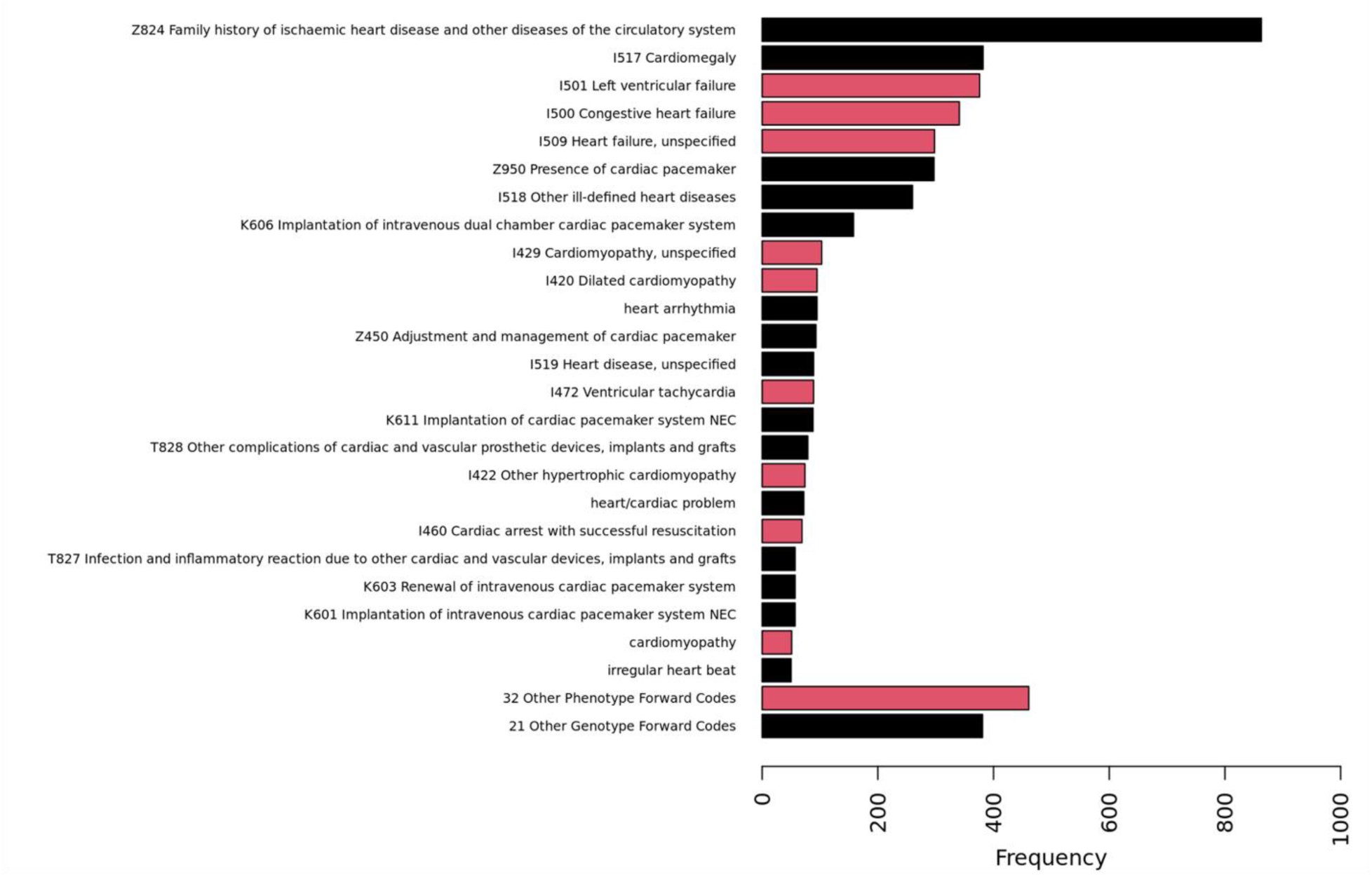
Cardiomyopathy code frequencies observed in pathogenic or likely pathogenic or predicted pathogenic variant of uncertain significance carriers. Black bars indicate criteria only included in genotype-forward disease classification; red bars indicate criteria included in both genotype- and phenotype-forward disease classification.

**Supplemental Figure 5.**
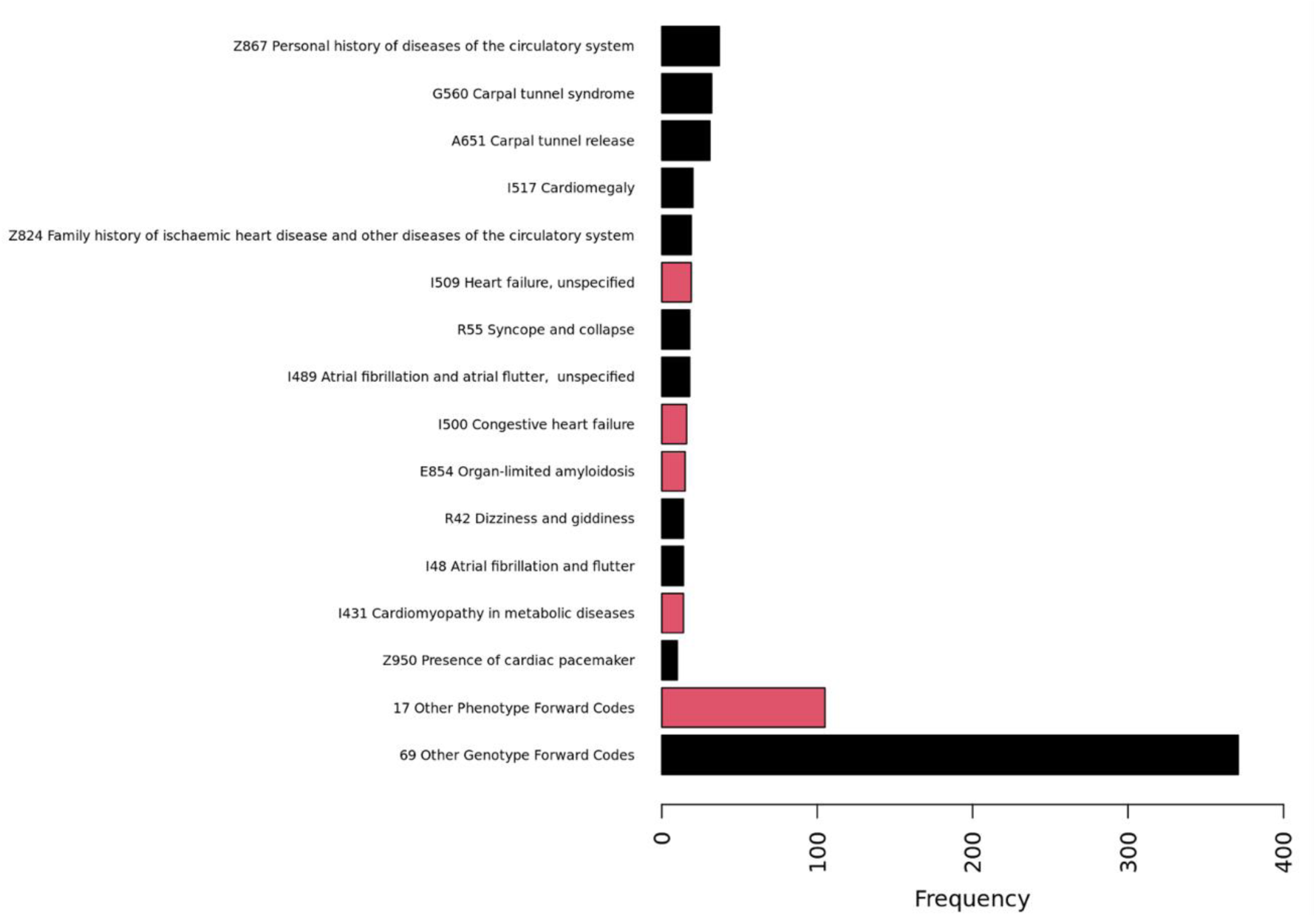
Hereditary transthyretin amyloidosis code frequencies observed in pathogenic or likely pathogenic or predicted pathogenic variant of uncertain significance carriers. Black bars indicate criteria only included in genotype-forward disease classification; red bars indicate criteria included in both genotype- and phenotype-forward disease classification.

**Supplemental Figure 6.**
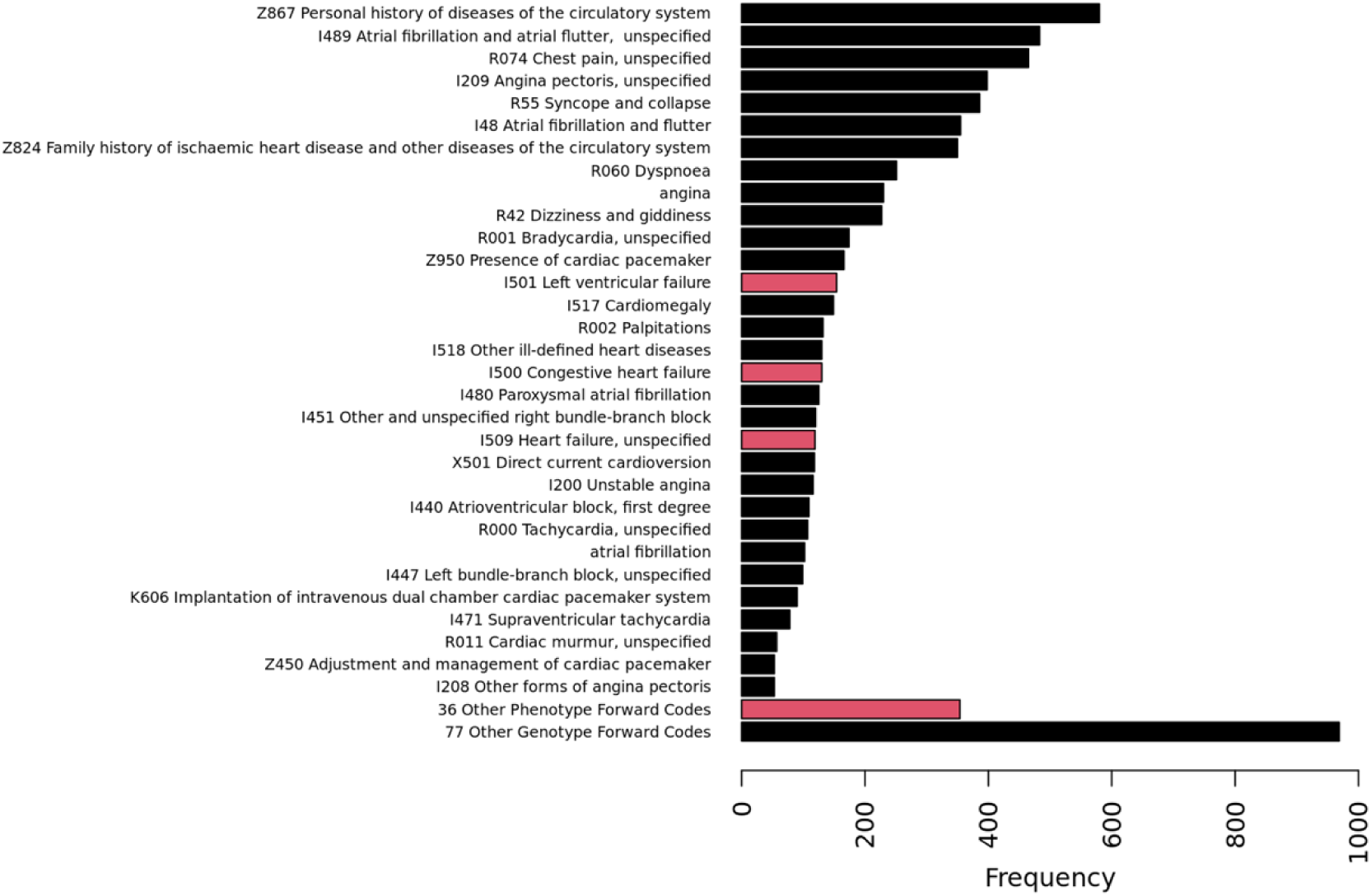
Arrhythmia disorder code frequencies observed in pathogenic or likely pathogenic or predicted pathogenic variant of uncertain significance carriers. Black bars indicate criteria only included in genotype-forward disease classification; red bars indicate criteria included in both genotype- and phenotype-forward disease classification.

**Supplemental Figure 7.**
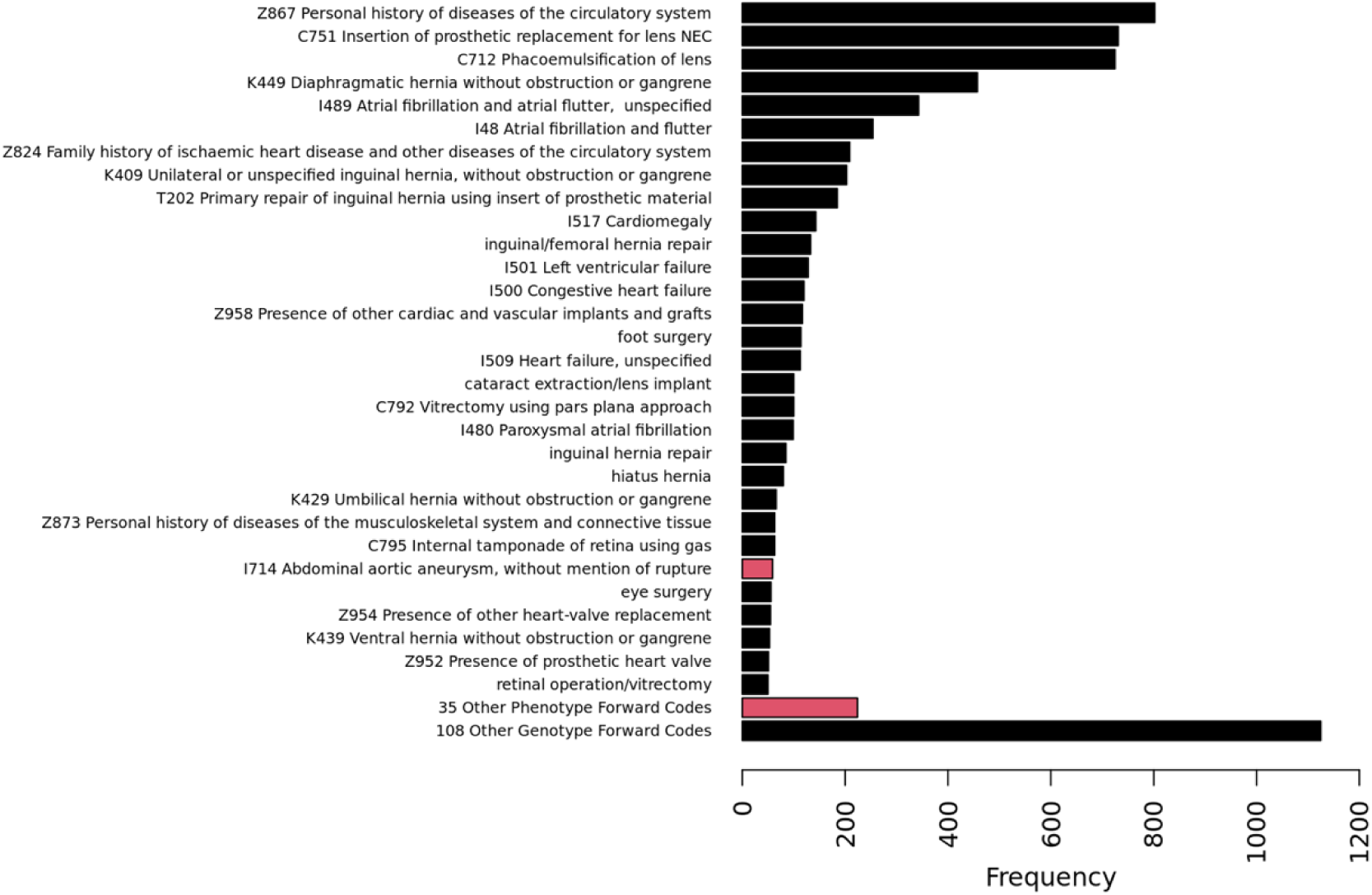
Aortopathy and connective tissue disorder code frequencies observed in pathogenic or likely pathogenic or predicted pathogenic variant of uncertain significance carriers. Black bars indicate criteria only included in genotype-forward disease classification; red bars indicate criteria included in both genotype- and phenotype-forward disease classification.

## Supplemental Tables

**Supplemental Table 1.** List of genes and associated monogenic cardiovascular disease.

**Supplemental Table 2.** Cardiomyopathy phenotype codes.

**Supplemental Table 3.** Hereditary transthyretin amyloidosis phenotype codes.

**Supplemental Table 4.** Familial hypercholesterolemia phenotype codes.

**Supplemental Table 5.** Aortopathy/connective tissue disorder Arrhythmia disorder phenotype codes.

**Supplemental Table 6.** Arrhythmia phenotype codes.

**Supplemental Table 7.** Genotype-forward pathogenic and likely pathogenic variant level carrier status frequencies with and without disease expression by monogenic cardiovascular disease genetic variant.

**Supplemental Table 8.** Summary of variants and disease expression in participants with two P/LP variants.

**Supplemental Table 9.** Genotype-forward predicted pathogenic VUS carrier status frequencies with and without disease expression by monogenic cardiovascular disease genetic variant.

**Supplemental Table 10.** Summary of variants and disease expression in participants with two pp-VUS.

**Supplemental Table 11.** Phenotype-forward disease expressive pathogenic and likely pathogenic carrier frequencies by monogenic cardiovascular disease gene.

**Supplemental Table 12.** Genotype-forward pathogenic variant of uncertain significance carrier status frequencies with and without disease expression by monogenic cardiovascular disease gene.

**Supplemental Table 13.** Phenotype-forward disease expressive pathogenic and likely pathogenic carrier frequencies by monogenic cardiovascular disease gene.

**Supplemental Table 14.** Phenotype-forward disease expressive predicted pathogenic variant of uncertain significance carrier frequencies by monogenic cardiovascular disease gene.

**Supplemental Table 15.** Phenotype criteria observed code frequencies for FH.

**Supplemental Table 16.** Phenotype criteria observed code frequencies for CM.

**Supplemental Table 17.** Phenotype criteria observed code frequencies for hTTR.

**Supplemental Table 18.** Phenotype criteria observed code frequencies for arrhythmia disorders.

**Supplemental Table 19.** Phenotype criteria observed code frequencies for aortopathies/connective tissue disorders.

